# Appropriately smoothing prevalence data to inform estimates of growth rate and reproduction number

**DOI:** 10.1101/2022.02.04.22270426

**Authors:** Oliver Eales, Kylie E. C. Ainslie, Caroline E. Walters, Haowei Wang, Christina Atchison, Deborah Ashby, Christl A. Donnelly, Graham Cooke, Wendy Barclay, Helen Ward, Ara Darzi, Paul Elliott, Steven Riley

## Abstract

The time-varying reproduction number (***R***_***t***_) can change rapidly over the course of a pandemic due to changing restrictions, behaviours, and levels of population immunity. Many methods exist that allow the estimation of ***R***_***t***_ from case data. However, these are not easily adapted to point prevalence data nor can they infer ***R***_***t***_ across periods of missing data. We developed a Bayesian P-spline model suitable for fitting to a wide range of epidemic time-series, including point-prevalence data. We demonstrate the utility of the model by fitting to periodic daily SARS-CoV-2 swab-positivity data in England from the first 7 rounds (May 2020 – December 2020) of the REal-time Assessment of Community Transmission-1 (REACT-1) study. Estimates of ***R***_***t***_ over the period of two subsequent rounds (6-8 weeks) and single rounds (2-3 weeks) inferred using the Bayesian P-spline model were broadly consistent with estimates from a simple exponential model, with overlapping credible intervals. However, there were sometimes substantial differences in point estimates. The Bayesian P-spline model was further able to infer changes in ***R***_***t***_ over shorter periods tracking a temporary increase above one during late-May 2020, a gradual increase in ***R***_***t***_ over the summer of 2020 as restrictions were eased, and a reduction in ***R***_***t***_ during England’ s second national lockdown followed by an increase as the Alpha variant surged. The model is robust against both under-fitting and over-fitting and is able to interpolate between periods of available data; it is a particularly versatile model when growth rate can change over small timescales, as in the current SARS-CoV-2 pandemic. This work highlights the importance of pairing robust methods with representative samples to track pandemics.

## 1 Introduction

Since the beginning of the COVID-19 pandemic, governments and policy makers have sought to strike a delicate balance between controlling the spread of the SARS-CoV-2 virus and allowing society to function as close to prepandemic levels as possible. Non-pharmaceutical interventions (NPIs) have often been introduced to curtail the spread of the virus by reducing rates of transmission, with varying levels of stringency from national lockdowns and curfews [1–3], to reduced opening hours for hospitality [4]. Despite their success in controlling the virus, NPIs have also been associated with negative impacts on the economy and on other facets of people’ s health, for example mental health [5]. Accurate and timely measurements of infection prevalence and its rate of growth are required to effectively inform individuals and governments so that interventions can be timely and proportionate.

Throughout the pandemic there has been a large amount of data collected in order to track the SARS-CoV-2 virus spread and prevalence of COVID-19 disease, with numerous data streams in England alone [6–10]. However, sources of pandemic data are prone to biases which may lead to effective sample populations unrepresentative of the underlying population. For example, mass testing, a bedrock of pandemic response, can be heavily influenced by the test supply, the propensity of individuals to seek tests and whether tests are only used on symptomatic individuals [11]. The REal-time Assessment of Community Transmission-1 (REACT-1) study [12] is a series of repeat crosssectional study that aims to accurately measure the community prevalence of SARS-CoV-2 in England over time. There has been a round of the study approximately monthly since May 2020 [6]. Due to the random sampling procedure it employs, it is relatively unbiased compared to many other sources of data collection, allowing accurate estimates of prevalence to be obtained over each round.

Prior pandemics [13] and standard epidemic theory suggest that prevalence should increase or decrease exponentially over substantial periods of time. For this reason, typical time-series analysis of epidemic data might include fitting an exponential model to the data under a constant growth rate assumption and from this inferring the reproduction number, *R* [14]. This is valid at small timescales, but due to depletion of susceptibles, NPIs, and changing behaviour over time, the assumption of constant growth rate is invalid over a longer duration. Various methods exist that allow the course of an epidemic to be studied over a longer period. The progression of the instantaneous reproduction number *R*_*t*_ over time has been estimated from incidence data with only assumptions of the serial interval required [15, 16]. These methods can have significant variation over small timescales [17] and so smoothing over time windows is often performed. However, this can make the models sensitive to assumptions on the degree of smoothing [18]. Even those methods that are able to avoid these limitations [19] are limited in their ability to deal with times-series data where a significant number of daily data is missing.

Splines are a versatile tool for modelling non-linear functions when the exact form of the function is unknown. However, underlying assumptions can lead to either over- or under-fitting. Penalised splines (P-splines) [20] seek to avoid over-fitting through the inclusion of discrete penalties on the basis coefficients, though this penalty has no exact interpretation in terms of the function’ s shape. Their Bayesian counterpart however [21] offers a statistically robust method of capturing variation in the data whilst also preventing overfitting through the inclusion of appropriate prior distributions that act on the functional form of the spline. Bayesian P-splines have been used to model the spatial variation in the risk of cholera infection [22], the effects of age, space and time on cancer mortality [23], and to model the relation between respiratory mortality and daily fine particle exposure [24]. Their use in epidemic time-series data however, has been limited [25], perhaps because major programming packages only allow for prior distributions that penalise changes in the main response function which would correspond to a prior distribution of constant prevalence which is problematic given expected exponential trends.

Here we develop a Bayesian P-spline model that may be widely applicable to epidemic time-series data. The model, in the absence of data, penalises changes in the growth rate, reflecting the prior knowledge we have on an epidemic system. We demonstrate the utility of the model by fitting to the daily swab-positivity data from the first 7 rounds of the REACT-1 study. The data is semi-continuous over time with well sampled periods of between 18 and 32 days for each round, with non-sampled periods between rounds.

## 2 Materials and methods

### 2.1 REACT-1 Data

The complete study protocol of REACT-1 has been described previously [26]. In brief, letters are sent randomly to named individuals from the list of GP patients in England held by the National Health Service, stratified by lower-tier local authorities (LTLAs, n=315). Those who agree to participate in the study self-administer throat and nose swabs (administered by parent or guardian for children aged 5-12 years). These are then collected and sent on a cold chain for analysis by reverse-transcript polymerase-chain-reaction (RT-PCR), with E- and N-gene targets, in a single commercial laboratory (round 1 also included some swabs tested in Public Health England laboratories). The REACT-1 study received research ethics approval from the South Central-Berkshire Research Ethics Committee (IRAS ID: 283787). The sample size of the study has ranged from 120,000 to 160,000 across the 7 rounds of data collection from May 2020 to December 2020 (Supplementary Table 1).

The cycle threshold (Ct) value of the E- and N-gene targets in the RT-PCR test is used as a proxy for intensity of the viral load (where Ct value is inversely proportional to viral load). A swab was defined as positive if 1) both the E- and N-gene targets were positive or 2) the N-gene target was positive with a Ct value less than 37. To test the sensitivity of our analyses we use different definitions of swab positivity: asymptomatic positives are defined as those who match our original positivity criteria but did not self-report having had any symptoms in the month prior to their swab test; double positives were defined as those with positive E- and N-gene targets; and lower threshold positives were defined in a similar way to our original positivity definition, using a Ct cut off value of 35 for the swabs with only N-gene target positive.

The date the swab test was completed and the date the test was collected were recorded. However, one or both dates are missing for some participants. In participants where both the date of swab test and date of collection are known, we observe that the two dates are highly consistent (86.9% are the same date, 9.6% are one day apart). Thus, we define a new composite variable that is equal to the date an individual completed the swab test, if available and within the range of dates for which they could plausibly have received the test (70.1% of individuals).

Participants provided information on the day they completed the swab test and the day it was collected is also recorded. However, one or both dates are missing for some participants. In participants where both the date of swab test and date of collection are known, we observe that the two dates are highly consistent (86.9% are the same date, 9.6% are one day apart). Thus, we define a new composite variable that is equal to the date an individual completed the swab test, if available and within the range of dates for which they could plausibly have received the test (70.1% of individuals), or, if the swab completion date is not available or is inconsistent, then we use the date of collection as a proxy (29.1% of individuals). For all rounds, over 90% of swabs had either a date of completion or a date of collection and at least 95% of positive swab tests had one of these dates associated (Supplementary Table 1).

### 2.2 Simple exponential model

As a baseline model to study how prevalence varies over short periods of time, we fit a simple exponential model with a constant growth rate:

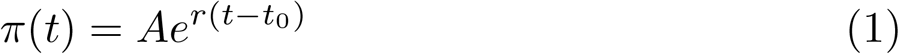

where *π*(*t*) is the prevalence at time *t, A* is the prevalence at time *t*_0_ (the first date in the data the model is fit to) and *r* is the growth rate. We fit the exponential model to multiple subsets of the data. First, to explore the rate of growth across the period covered by two rounds, we fit the model to the data from each pair of subsequent rounds. Secondly, we explore within round growth rates by fitting the model to the 7 individual rounds of data.

The average reproduction number for the period described by each model was estimated assuming a generation time following a gamma distribution with shape parameter *α* = 2.29 and rate parameter *β* = 0.36 [27]. The reproduction number was then calculated from the estimate of the growth rate [28], *r*, using the equation:

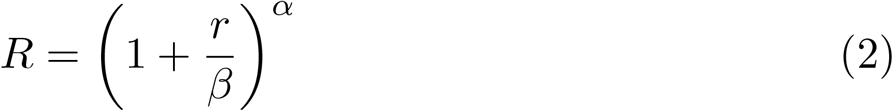

which is valid for *r >* −*β*.

### 2.3 Bayesian P-spline model

#### 2.3.1 The Model

A general model for prevalence using smoothed functions can be written as

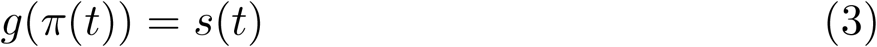

where *π*(*t*) is, as before, the prevalence at time *t, s*(*t*) is the value of a general smooth function at time *t*, and *g* is the link function, which we take as the logit function for the REACT-1 binomial data. We define our smooth function as a linear combination of *N* B-splines (see below) of order *n*:

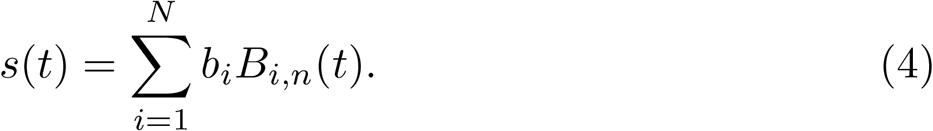

The B-splines are defined by a non-decreasing sequence of knots *t*_1_ … *t*_*q*_ and the polynomial degree, *p* (*n* − 1). The first order (*p* = 0) B-splines are defined as

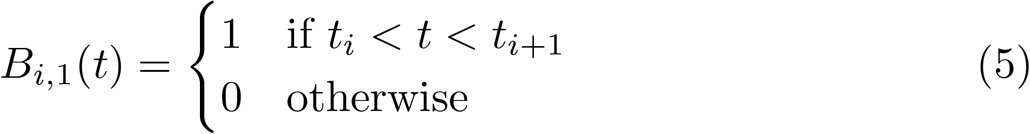

and then higher order B-splines are defined recursively

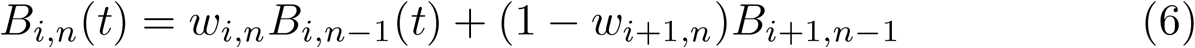

where,

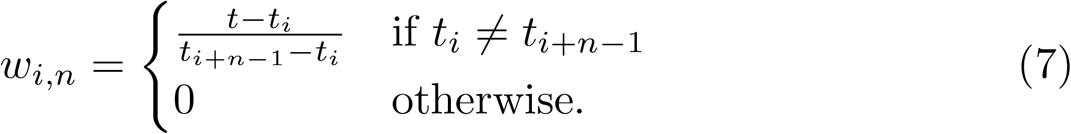

To model how prevalence varies over time we define a family of 4^*th*^ order Bspline functions over the duration of the study. The locations of knots are placed at regular intervals over the entire duration of the study so that there are approximately 5 days between each pair of adjacent knots (approximately as the length of data is not always divisible by 5). Additionally, the model is extended a further 3 knots from both the beginning and end of the study so that the model output for the period of interest is not affected by the irregular B-splines defined at the boundaries. These extensions of the model outside of the duration of the study are not included in any visualisation after fitting. Models using different knot sizes were investigated, and a knot size of 5 days was chosen for the model used in the main analyses. The knot size of 5 days was chosen to ensure that the model was not under-fitted, whilst also not allowing the fitting of the model to become too computationally expensive.

In order to limit over-fitting of the model to noise in the data, we define a second-order random-walk prior distribution [21] on the coefficients:

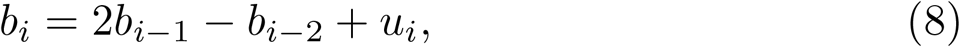

where

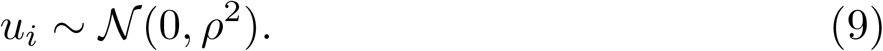

This prior distribution penalises any change from the first d erivative o f the smoothing function, which, for a logit link function, is approximately equal to the growth rate at low prevalence, and for a log link function is exactly equal to the growth rate. The amount to which changes of the first derivative of the smooth function are penalised is controlled by another parameter of the model, *ρ*, that we give a loose but proper inverse gamma prior distribution *ρ*∼ℐ 𝒢 (0.001, 0.001). The first t wo c oefficients of the mod el, *b*_1_ and *b*_2_, are given an uninformative constant prior distribution. A first-order random-walk prior distribution was also considered: *b*_*i*_ = *b*_*i*−1_ + *u*_*i*_, where *u*_*i*_ is as before. However, a comparison of both models fit to simulated data of an exponentially increasing prevalence (Fig. 1) showed that a first-order random-walk prior distribution, in the absence of data, favours a constant prevalence, which is unnatural for an epidemic system.

**Fig. 1.**
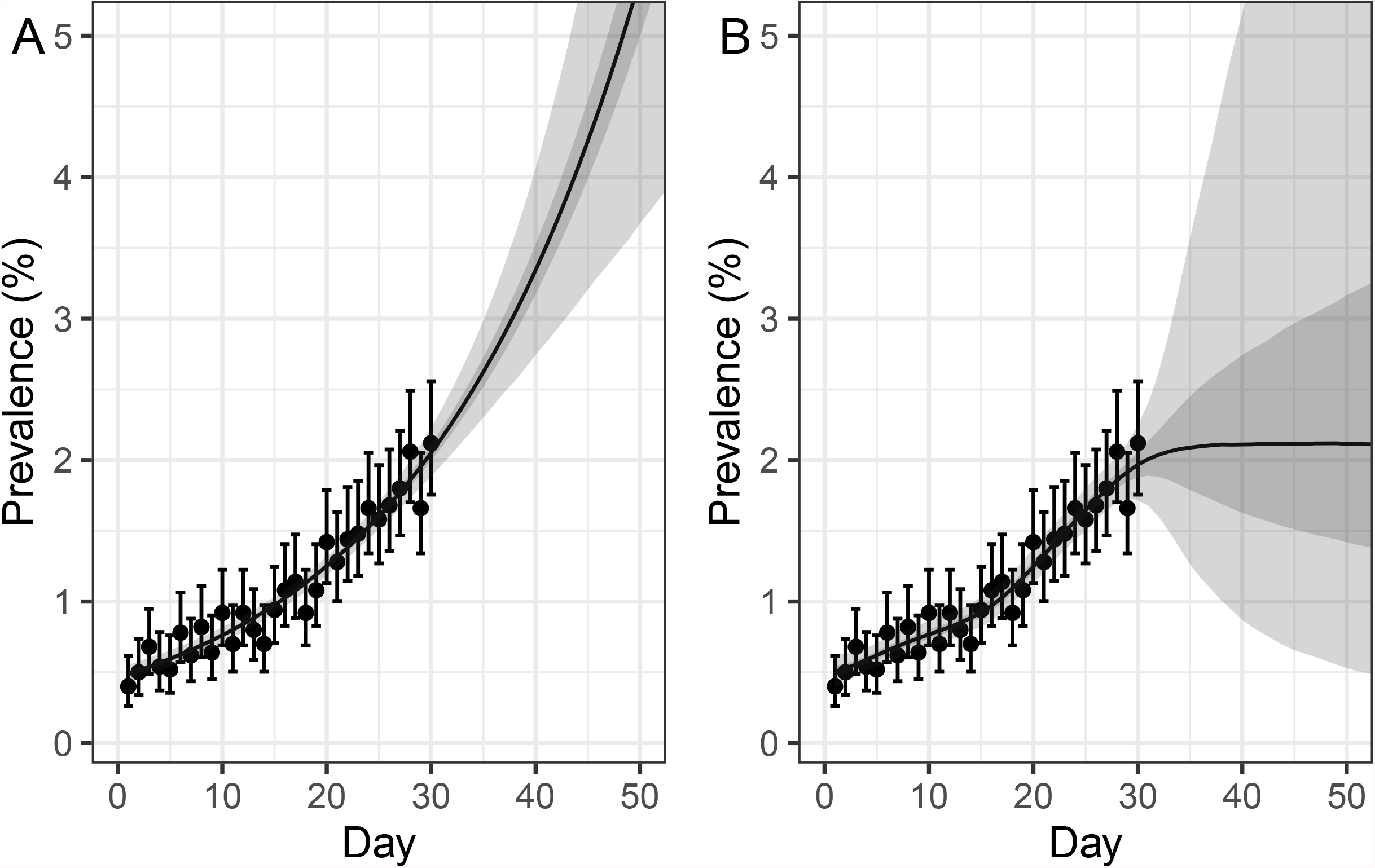
Comparison of Bayesian P-spline models with second-order and first order random-walk prior distributions. Simulated Binomial data for an exponentially increasing prevalence, *π*(*t*) = 0.5 *× exp*(0.05 *× t*), with 5000 tests performed a day. Daily estimates of prevalence (points) and 95% confidence intervals are shown. (A) Model fit for a Bayesian P-spline model with a second-order random-walk prior distribution. (B) Model fit for a Bayesian P-spline model with a first order random-walk prior distribution. (A, B) Central estimates (solid line) are shown with 50% (dark shaded region) and 95% (light shaded region) credible intervals are also shown. Models have been defined up to 60 days, despite the data only going up to 30 days, in order to demonstrate the effect of the model’ s prior distribution during periods of no data.

The model is implemented in STAN and parameter posteriors are sampled using a No-U-Turn Sampler [29] assuming a Binomial likelihood. Four chains are run and the performance of the model fitting is checked by looking at measures of the Bulk LSS and individual parameters’ potential scale reduction statistic, 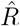. An analogous Bayesian P-spline model is also fitted to Public Health England Pillar 1 and 2 case (by specimen date) data [10]. Instead of prevalence, the model fits to the daily number of cases using a log link function. The model is fitted assuming a Negative-Binomial likelihood, with the additional over-dispersion parameter that is required given an uninformative constant prior distribution.

When fitting the model to different subsets of the data, despite the target knot size being 5 days the actual knot size was not always exactly 5 days. In order to compare the estimated value of *ρ* between models using slightly different knot sizes we defined the standardized 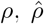. The value of *ρ* should increase linearly with knot size for small knot sizes (for large knot sizes linearity will break down due to under-fitting). Therefore, we defined 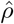 as the value that *ρ* would have for a knot size of exactly 5 days assuming linearity.

#### 2.3.2 Calculating average growth rate over a finite period

In order to compare the fit of the Bayesian P-spline model to the more simple exponential model it is necessary to calculate the average growth rate over finite periods of time of the model output. These can then be compared to the average growth rates calculated for the same periods of time by the exponential model. For any set of sampled parameters from the posterior we can calculate the prevalence at any time during the study period. We then assume that the prevalence at a time *t*_2_ is linked to the prevalence at time *t*_1_ by a constant growth rate 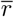 such that

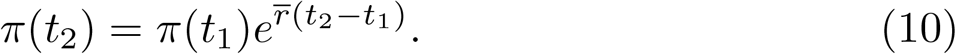

Then we can calculate the average growth rate between two times for any set of parameters sampled from the posterior through the equation:

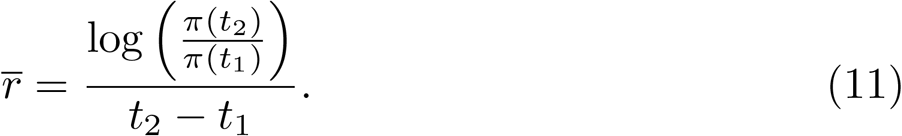

By calculating this value over the entire posterior of parameter values we can calculate the posterior probability for the average growth rate between two time points of our model. This average growth rate can then be converted into an estimate for the average reproduction number over the period using equation 2. These estimates are analogous to the previous estimates found for the exponential model and so can readily be compared.

#### 2.3.3 Calculating instantaneous growth rate

The rate of change of prevalence can be written in terms of a time varying growth rate, *r*(*t*)

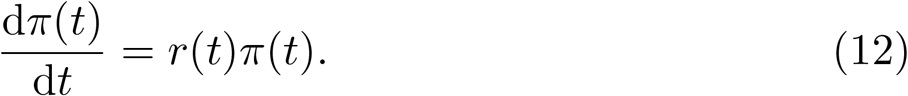

This then has the solution,

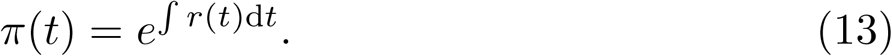

Equating the two equations (equations 3 and 13) for prevalence, *π*(*t*) we have

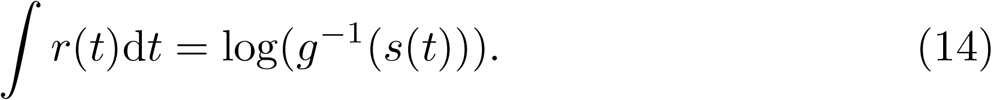

This can be solved for the instantaneous growth rate, *r*(*t*), through differentiation with respect to time and Leibniz’ s rule giving:

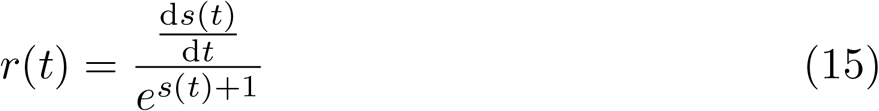

for the case in which *g* is the logit function. The smooth function, *s*(*t*) and its first derivative with respect to time are defined over the entire period of the study. We calculate *r*(*t*) over the duration of the study for all combinations of parameters in our sampled posterior. This gives us the posterior distribution of *r*(*t*) for the entire study period. For the case in which *g* is the log function *r*(*t*) is simply the first derivative with respect to time of *s*(*t*).

#### 2.3.4 Calculating reproduction number from prevalence estimates

If the rate of secondary infections at time *τ* since infection is given by *η*(*τ*) then we can write the reproduction number as

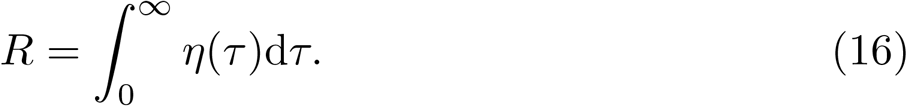

Normalising *η*(*τ*) gives us the generation time *g*(*τ*)

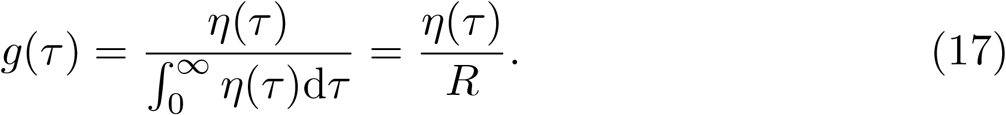

Writing the prevalence at time *t, π*(*t*), in terms of the prevalence at times less than *t* we get,

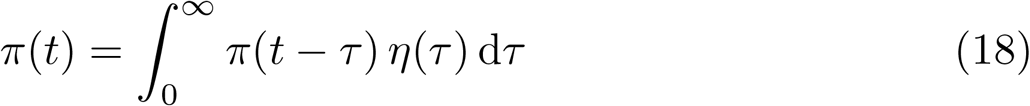

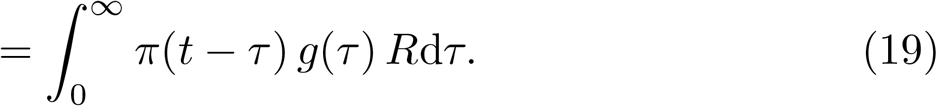

If we make the assumption that *R* has been constant for a significant period of time before *t* then we can rearrange the expression [28] to get

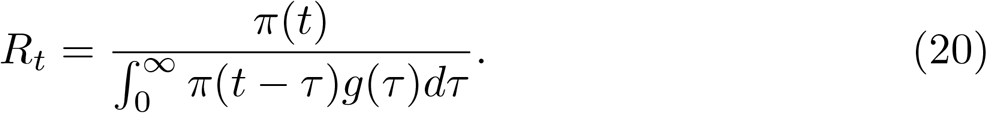

The fit of the Bayesian P-spline model gives us the posterior distribution of *π*(*t*) for the entire duration of the study. We again take the generation time *g*(*τ*) to be an inverse gamma distribution with scale parameter 2.29, and rate parameter 0.36 [27]. The rolling 14 day average of *R*_*t*_ can then be calculated from the above equation by integrating over the previous 14 days at each point in time. However, this calculation relies on the approximation that *R*_*t*_ is constant over the 14 days. Note that 14 days was chosen so that the generation time distribution had declined to a negligible amount. By calculating *R*_*t*_ for the entire posterior of our prevalence estimates we can obtain an appropriate confidence interval for our estimates of *R*_*t*_.

## 3 Results

### 3.1 Comparison of exponential model and Bayesian P-spline model

The growth rates of prevalence between rounds were assessed by fitting the simple exponential model to each pair of subsequent rounds (Fig. 2a). The Bayesian P-spline model was fitted to all 7 rounds (Fig. 2c) and, from this continuous estimate of prevalence, estimates of the average growth rates over the same subsequent rounds were also obtained. The Bayesian P-spline model was also fit to an increasing number of rounds (from round 1 only to all 7 rounds) and the average growth rate for the final two rounds of each fit was estimated. Estimates of *R* and doubling/halving times were then calculated from all estimated growth rates (Supplementary Table 2). Good agreement was found between estimates obtained from the simple exponential models and those obtained from the Bayesian P-spline models with overlapping credible intervals (Fig. 3a). The estimates obtained for rounds 6 and 7 showed the greatest level of disagreement with *R* estimated at 0.94 (0.92, 0.95) for the exponential model, but at 0.99 (0.95, 1.04) for the Bayesian P-spline model.

**Fig. 2.**
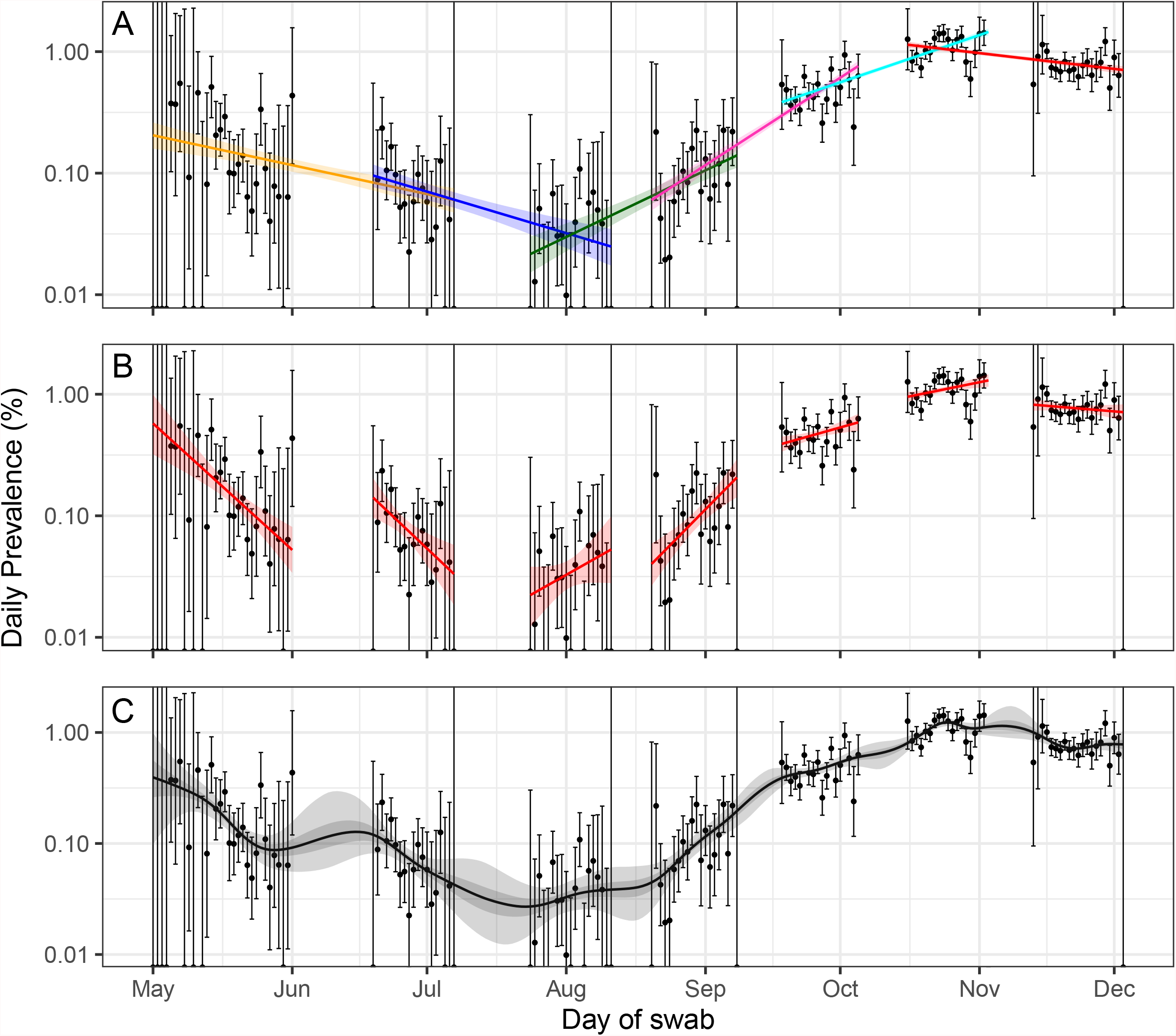
Exponential and Bayesian P-spline model fits. A: Exponential models fit to rounds 1 and 2 (yellow), rounds 2 and 3 (blue), rounds 3 and 4 (green), rounds 4 and 5 (pink), rounds 5 and 6 (cyan), and rounds 6 and 7 (red) of REACT-1 with shaded regions showing the 95% credible intervals. B: Exponential models fit to individual rounds (red) of REACT-1 with shaded regions showing the 95% credible intervals. C: Fit of the Bayesian P-spline model to all 7 rounds of REACT-1. Shaded regions show central 50% (dark grey) and 95% (light grey) credible intervals. Daily prevalence estimates (points) are shown with 95% confidence intervals (error bars).

**Fig. 3.**
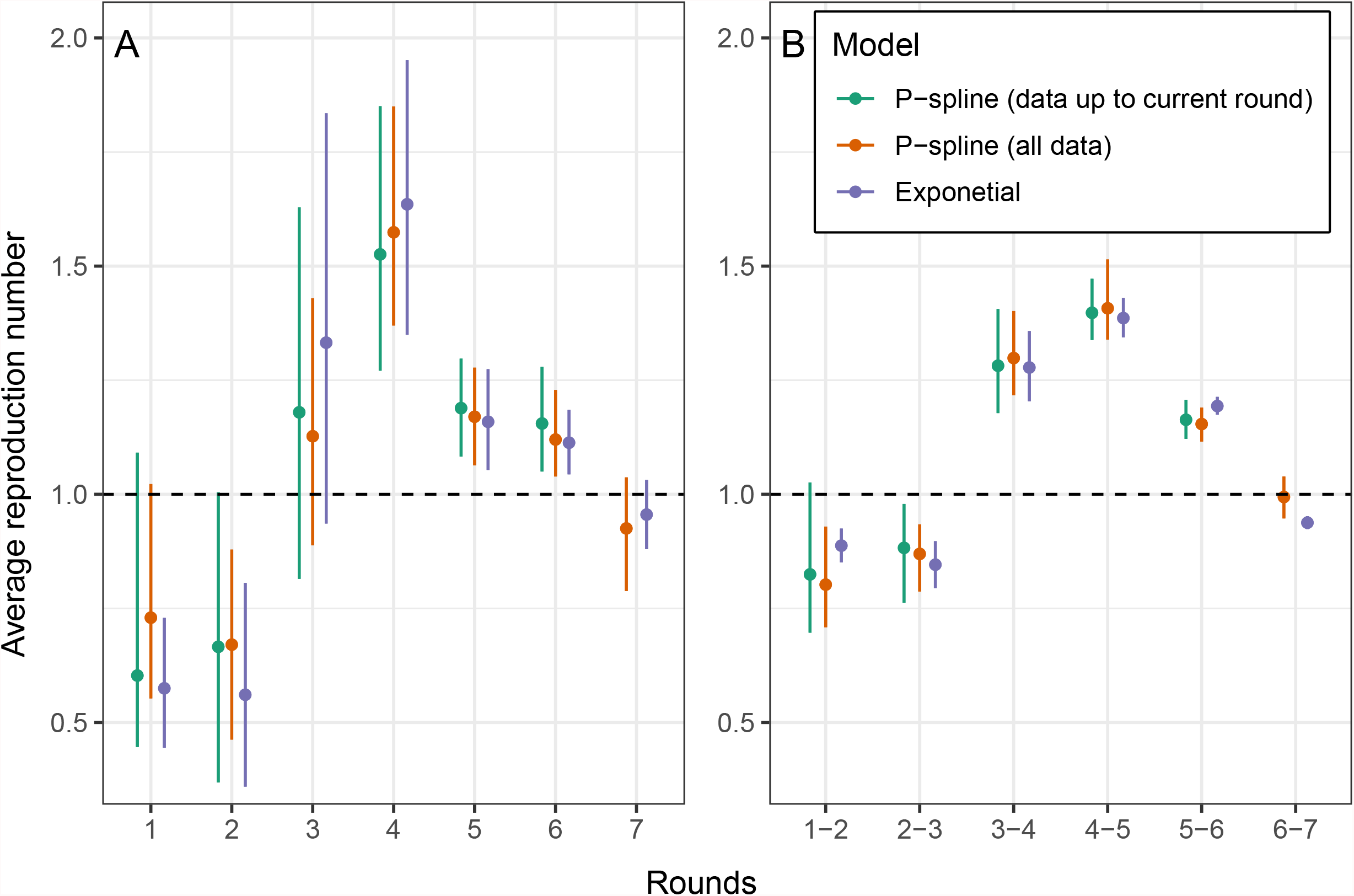
Estimated *R* for fixed periods. Comparison of *R* calculated for fixed periods using the exponential model (purple), and the Bayesian P-spline model fit using the first 7 rounds of REACT-1 (orange) and using all rounds up to the round for which *R* is being estimated (green). A: Estimates over the period of pairs of subsequent rounds. B: Estimates over the period of each individual round.

Similarly estimates of growth rates, *R*, and doubling/halving times were estimated for each individual round using the simple exponential model (Fig. 2b), the Bayesian P-spline model, and the Bayesian P-spline model fit to an increasing number of rounds (Supplementary Table 2). As before, there was good agreement between the estimated values of *R* with overlapping credible intervals for all rounds (Fig. 3b). However, there were clear differences in the posterior distributions of the estimates and their median values. Estimates of *R* obtained from the Bayesian P-spline model (fit to all 7 rounds) were higher than corresponding estimates from the exponential model over the periods of round 1 and round 2. During the periods of rounds 3 and 4 *R* estimates from the exponential model were higher than that of the Bayesian P-spline. There was a greater level of agreement between the two estimates for the periods of round 5, round 6 and round 7. Estimates of *R* obtained from the Bayesian P-spline model fit to subsets of the data (including all rounds up to the round for which *R* was estimated) showed similar point estimates and probability density to the Bayesian P-spline model fit to all data, with the exception of round 1 where it was closer in estimate to the exponential model. This was most likely due to the greater level of uncertainty at the beginning of round 1, and the fact that not enough data had been included to accurately estimate the parameter *ρ* which controls the degree to which the growth rate changes over time.

### 3.2 Continuous estimates of growth rate and R

Instantaneous growth rate was estimated from the prevalence estimates of the Bayesian P-spline model (Fig. 4). Changes in growth rate at a time frame less than the duration of a round (a few weeks) were detected as well as changes in growth rate in the periods between rounds. There was a significant level of variation in growth rate during the periods of round 1 (1 May - 1 June 2020) and round 2 (19 June - 7 July 2020), a period over which England’ s lockdown restrictions were just beginning to ease. In the period between rounds 1 and 2 the instantaneous growth rate temporally became positive, indicative of a growing epidemic. The growth rate was approximately constant during the periods of round 3 (24 July - 11 August 2020), 4 (20 August - 8 September 2020) and 5 (18 September - 5 October 2020), but in between these rounds there was a significant level of change with the growth rate increasing from round 3 to 4 and then decreasing significantly into round 5. During the period after round 5 to the end of round 7 (3 December 2020) there was significant variation in growth rate with it increasing above 0 and decreasing below 0 three times. We were also able to estimate the date at which prevalence began to increase, heralding the second wave of the pandemic. The day of minimum prevalence (Fig. 5) was found to be 23 July 2020 (13 July 2020, 16 August 2020).

**Fig. 4.**
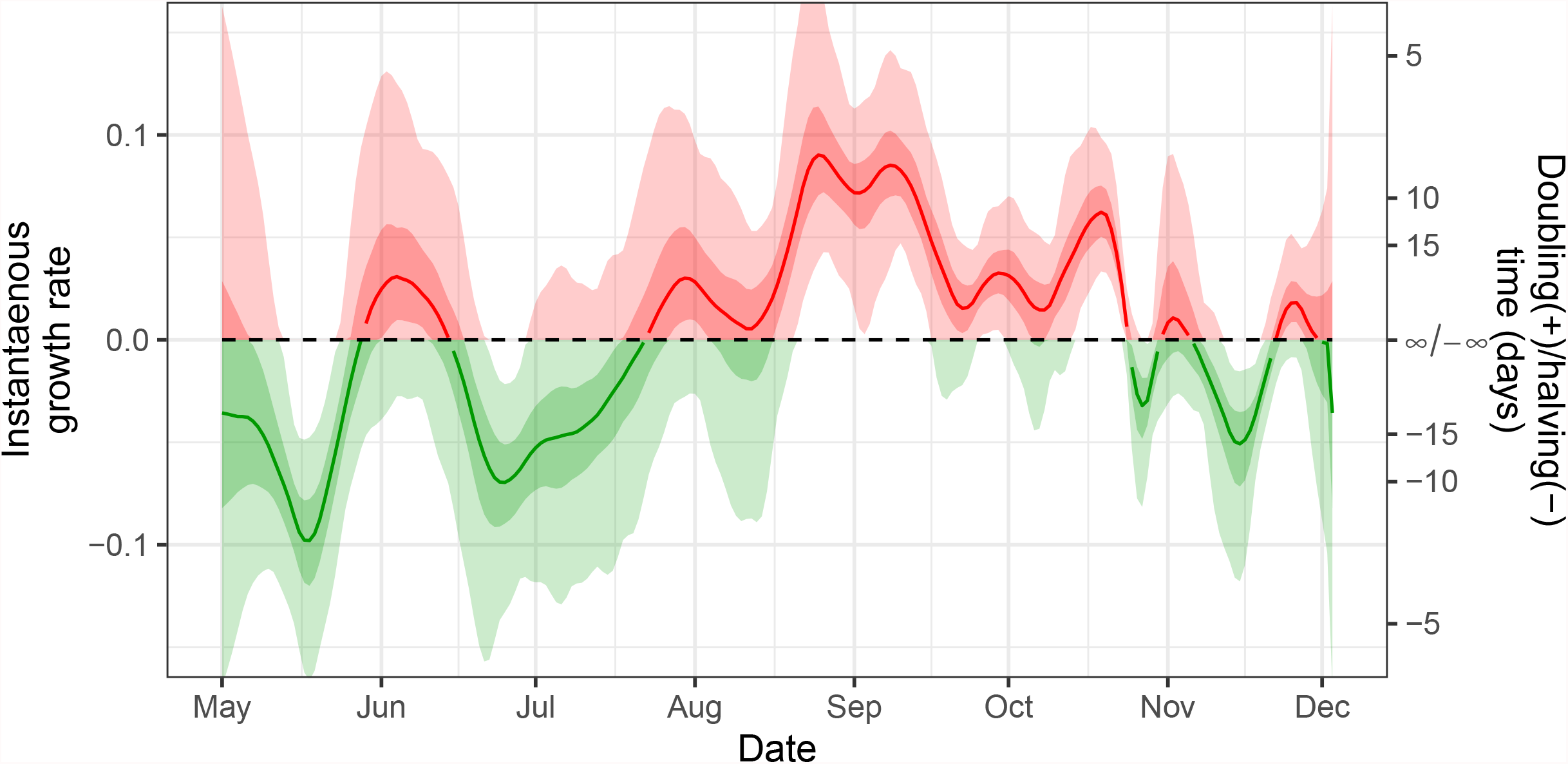
Instantaneous growth rate. The instantaneous growth rate over the study period as inferred from the Bayesian P-spline model. The Y axis on the right shows the corresponding doubling/ halving time corresponding to the growth rate on the left Y axis. The dotted line shows where growth rate = 0 and so the point of transition between epidemic growth and decline. The light shaded regions show the 95% credible interval and the dark shaded regions show the 50 % credible interval. Red highlights regions of the credible interval with values of growth rate greater than 0. Green highlights regions of the credible interval with values of growth rate less than 0.

**Fig. 5.**
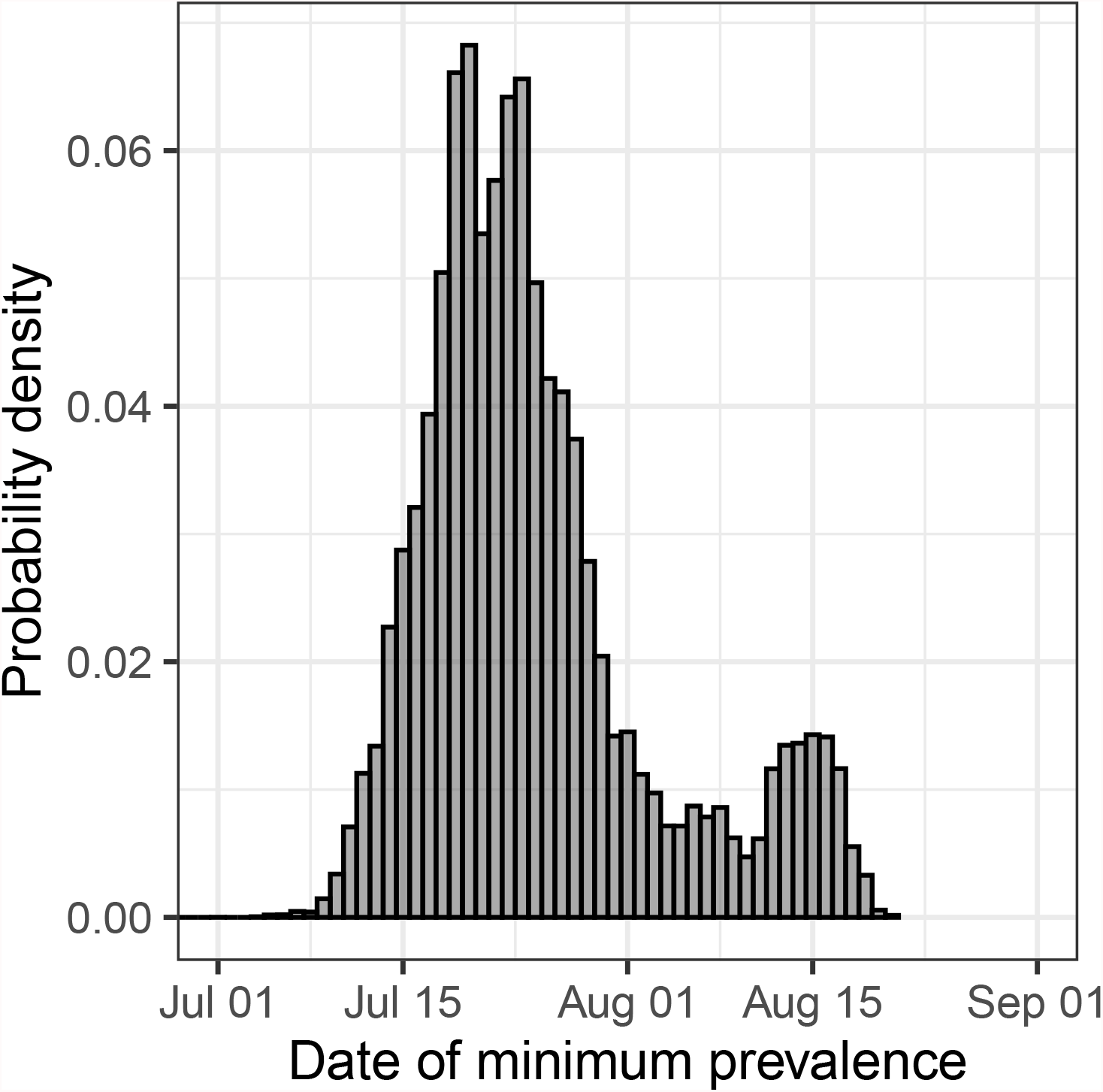
Date of minimum prevalence. Posterior probability distribution for the date at which prevalence was at its lowest in England from 1 May 2020 to 3 December 2020 estimated from the Bayesian P-spline model.

Previously we estimated the reproduction number from the Bayesian P-spline model through a direct conversion of the average growth rate for a fixed period of time. However, using the continuous estimates for prevalence we were able to use a more appropriate method (see Methods) to calculate the rolling two week average instantaneous reproduction number, *R*_*t*_, for the entire study (Fig. 6). As expected *R*_*t*_ behaved similarly to the instantaneous growth rate over the study period. There was a brief period in late-May/early-June 2020 in which *R*_*t*_ increased above 1 before decreasing below 1 going into late June 2020. *R*_*t*_ then once again increased, becoming greater than 1 towards the end of August 2020 where it remained until late October 2020 when it temporarily decreased below 1 until the end of November 2020. At the end of the study period (3 December 2020) it had a value of 1.03 (0.76, 1.29) with a 58% probability of being greater than 1.

**Fig. 6.**
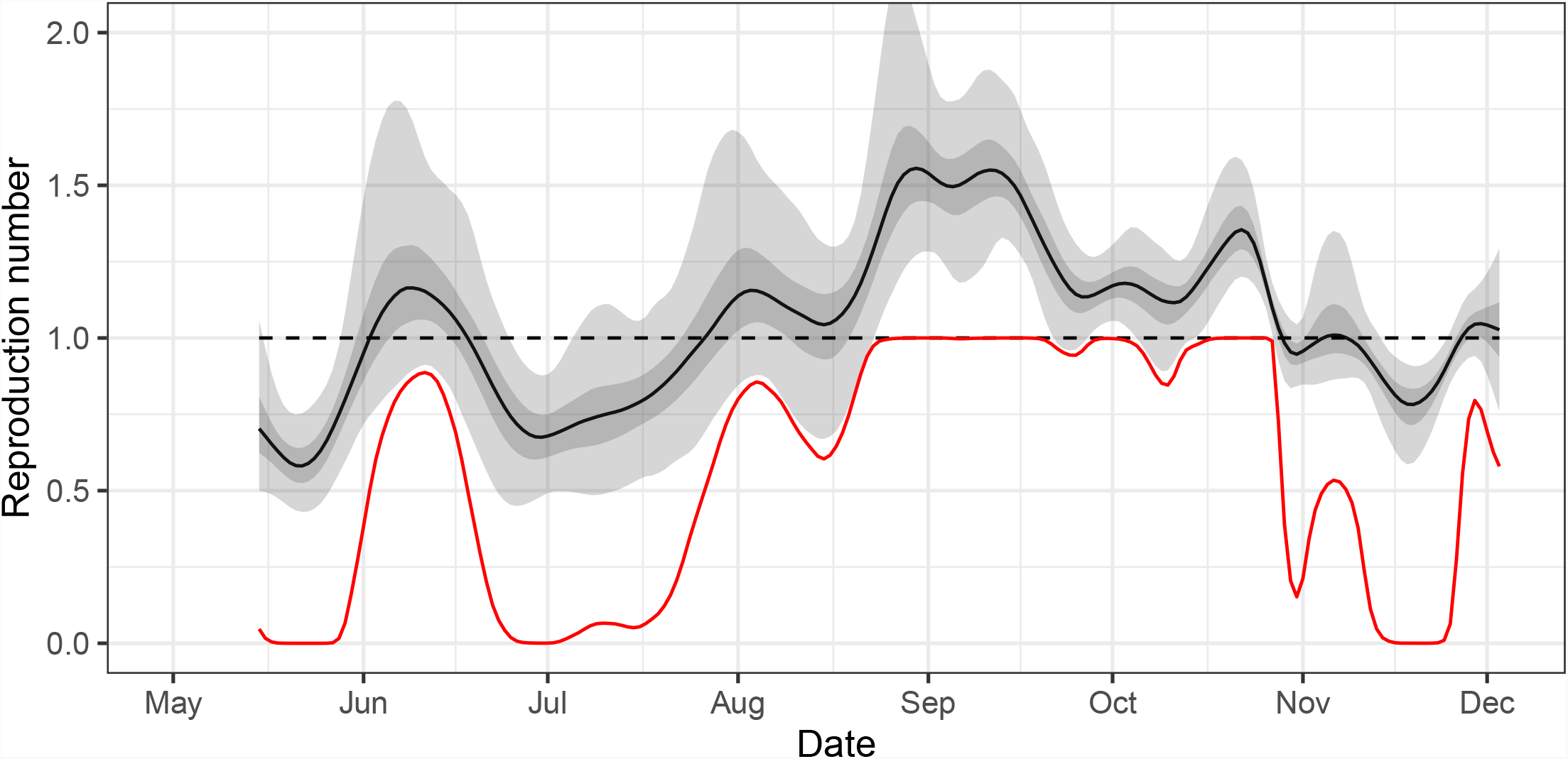
Continuous estimate of Reproduction number. The rolling two week average instantaneous reproduction number, *R*_*t*_, over the duration of the study calculated from the prevalence estimates of the Bayesian P-spline model. The central estimate is shown (solid black line) with 50% (dark grey shaded region) and 95% (light grey shaded region) credible intervals. The dotted line shows where *R*_*t*_ = 1 and so the point of transition between epidemic growth and decline. The red line shows the probability that *R*_*t*_ is greater than 1 over time.

Sensitivity analyses were performed by fitting the Bayesian P-spline model to subsets of the positive samples. Models were fit to subsets of data only including positive samples in which: the individuals did not report having symptoms (asymptomatic individuals), both gene targets were detected as positive, and where samples would still be defined as positive using a lower N-gene Ct value of 35 to define positivity. The model estimates for growth rate and *R*_*t*_ exhibited mainly similar patterns to the model fit using all positive samples (Supplementary Figures 1 and 2), with any divergences having overlapping credible intervals. Modelled estimates of growth rate and *R*_*t*_ obtained from fitting to positives defined with a more stringent definition of positivity (N-gene Ct value less than 35), showed far less variation over time, though exhibited broadly similar patterns.

### 3.3 Assessing presence of under-fitting

The second-order random-walk prior distribution used in the Bayesian Pspline model acts to smooth the estimates of prevalence over time and thus limit over-fitting. However, if the estimates are smoothed too heavily then statistically significant features of the data will not be fitted to. This underfitting of the Bayesian P-spline model was assessed by fitting the model multiple times with different knot sizes. The model parameter *ρ* that controls the second-order random-walk prior distribution should, if there is no under-fitting, increase linearly (as should its standard deviation) as the knot size increases (distance between knots increases linearly). At small knot sizes the value of *ρ* and its standard deviation increases linearly with the size of knots used (Fig. 7). However, for knot sizes larger than approximately 6 days the linearity of the relationship breaks down. The linear section of the graph included the knot size of 5 days which we have used for the main analysis and so no under-fitting was found to have occurred. The fit of the model for knot sizes of 2.5, 5 and 10 days were also inspected visually; no drastic difference was seen for the models with knot sizes of 2.5 and 5 days, but the model with a knot size of 10 days showed less variation in prevalence over time especially from late-October to mid-November 2020 most likely due to being in the under-fitting regime (Supplementary Figure 3).

**Fig. 7.**
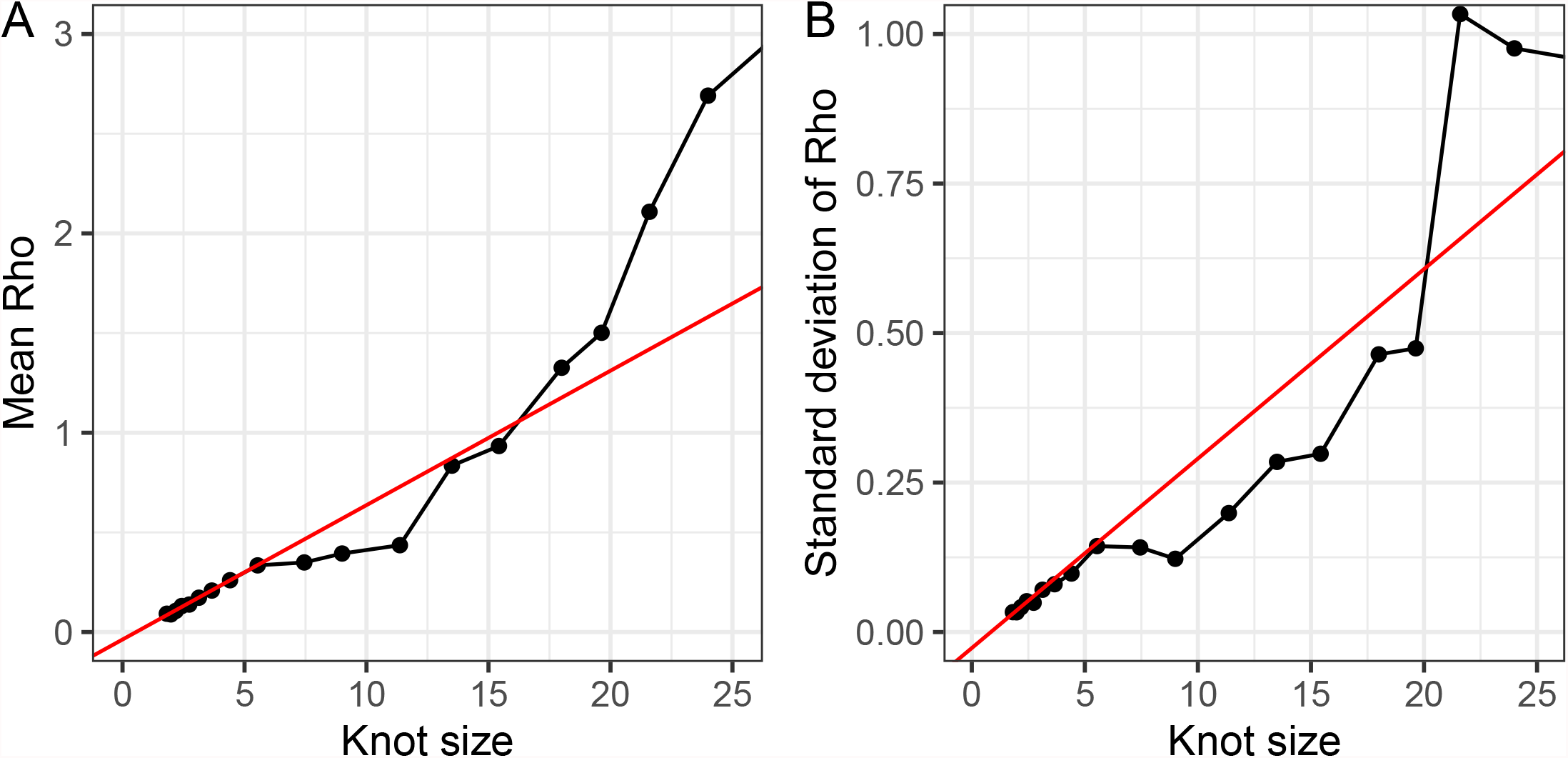
Effect of knot size on the parameter. *ρ*. The estimated value of the parameter *ρ* and its standard deviation from models using different knot sizes. The x-axis shows the knot size used in each model fit, the smaller the value the greater the number/density of knots. The red line shows a best fit line to the first four points in each graph and reflects the parameter values that would be expected if the model did not under fit when at a low density of knots.

### 3.4 Effect of increasing data

We investigated how the fit of the Bayesian P-spline model changed as data over a longer period of time was included (Fig. 8). Inclusion of new data can not only influence the fit of the model at the most recent points in time, but through changing the estimate of the parameter *ρ* can influence the model output over the entire period. The fit of the model to just round 1 of the data was remarkably close to an exponential model, though with larger uncertainty towards the beginning of the round. When round 2 was included the model fit to round 1 changed substantially, with a flatter prevalence for the first two weeks of round 1. This was due to the much larger estimate of the standardized 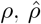, (increasing from 0.23 to 0.51). As rounds 3, 4 and 5 were included in the model fitting, the estimated value of 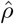 decreased, returning to a value of 0.23 with the inclusion of 5 rounds. When round 6 was included 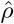 again increased due to clear variation in prevalence over a small timescale (a fall and rise were observed within the period of round 6), but when the final round, round 7, was included 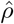 once more decreased. The effect of 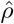decreasing is most easily observed in the fit of the model at the beginning of round 1 (going back to being closer to exponential) and between round 1 and round 2 (shrinking uncertainty in the increase in prevalence during the period).

**Fig. 8.**
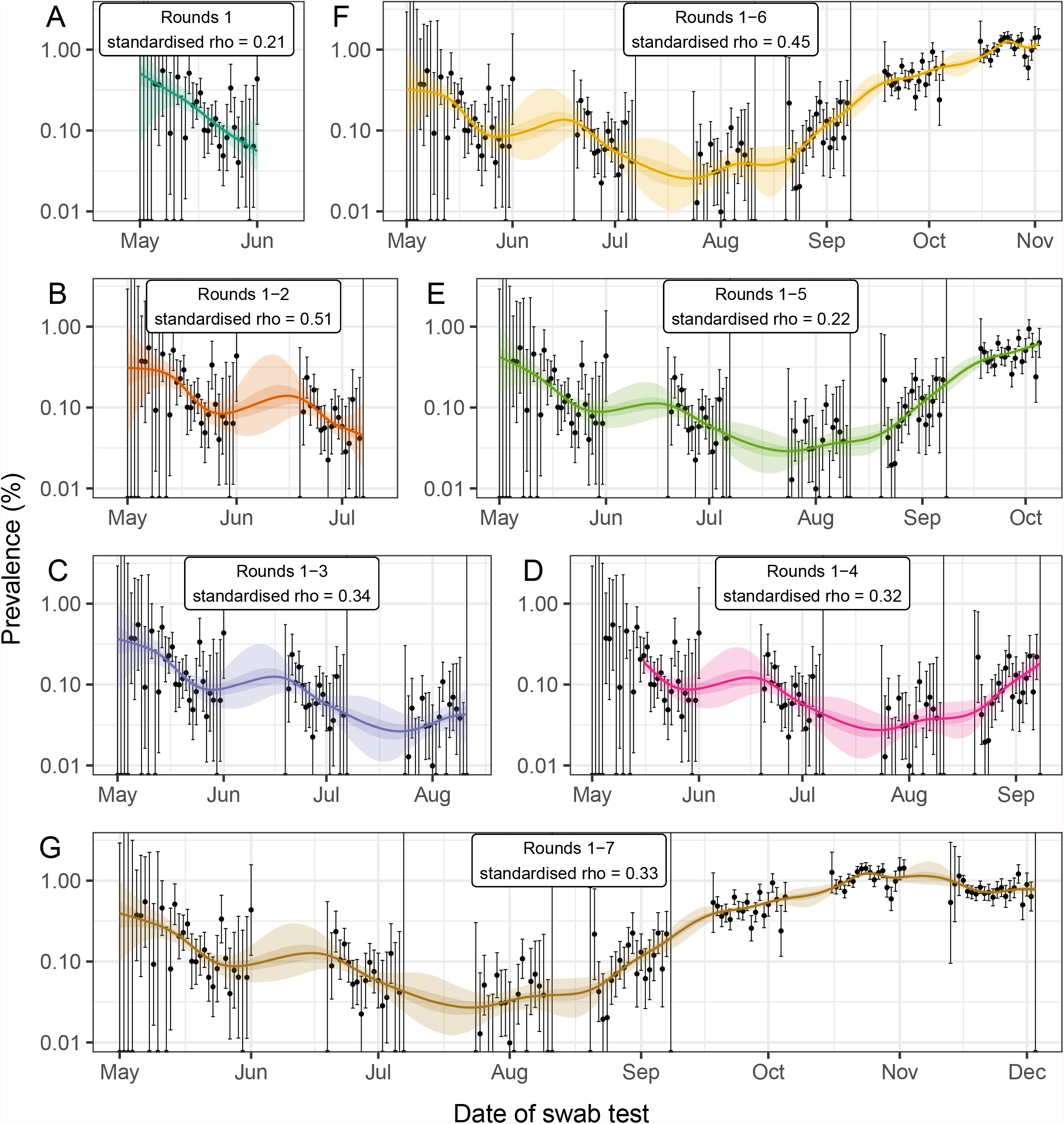
Comparison of models fitted to subsets of the data. Bayesian P-spline model prevalence estimates for the REACT-1 data as each new round is included in the model. (A) Model fit to round 1 only (dark green). (B) model fit to rounds 1 and 2 (orange). (C) Model fit to rounds 1 to 3 (purple). (D) Model fit to rounds 1 to 4 (pink). (E) Model fit to rounds 1 to 5 (light green). (F) Model fit to rounds 1 to 6 (yellow). (G) Model fit to rounds 1 to 7 (brown). The standardised *ρ* value is given for each model and is standardised for a knot size of 5 days (the target knot size for each model). Central model estimate is shown (solid line) with central 50 % (dark shaded region) and 95 % (light shaded region) credible intervals. Daily estimates of prevalence (points) are shown with 95% confidence intervals (error bars).

### 3.5 Model fit to public case data

In order to compare the trends in publicly available case data with REACT-1 prevalence a Bayesian P-spline model was fit to the Public Health England Pillar 1 and Pillar 2 case (by specimen date) data [10] allowing smoothed estimates of the daily number of cases to be obtained (Fig. 9). Broadly similar patterns to the modelled prevalence of REACT-1 were observed, but some differences were detected. The expected number of cases decreased steadily from early May 2020 reaching a minimum on 7 July (30 June, 16 July) 2020, earlier than the date in which prevalence as measured by REACT-1 reached a minimum (though with overlapping credible intervals). Further, estimates of instantaneous growth rate and *R*_*t*_ inferred from the model fit to case data showed far less variation over time than their REACT-1 model counterparts. Similarly to before, a small uptick in the growth rate was observed in June 2020, but the increase was not statistically significant and did not go above zero, the threshold for epidemic growth. The estimated *R*_*t*_ was below 1 until early July 2020; it then remained greater than 1 until early November 2020, plateauing at approximately 1.35 for the length of September 2020. *R*_*t*_ decreased steadily during the month of October 2020 in contrast to the trends observed in REACT-1 which measured a rapid decrease in *R*_*t*_ in midSeptember 2020 and a temporary rise in mid-October 2020. During November and early December 2020 *R*_*t*_, as measured from the case data, remained below 1, with no suggestion of an increasing epidemic on 3 December 2020 as was the case for the model fit to REACT-1 data.

**Fig. 9.**
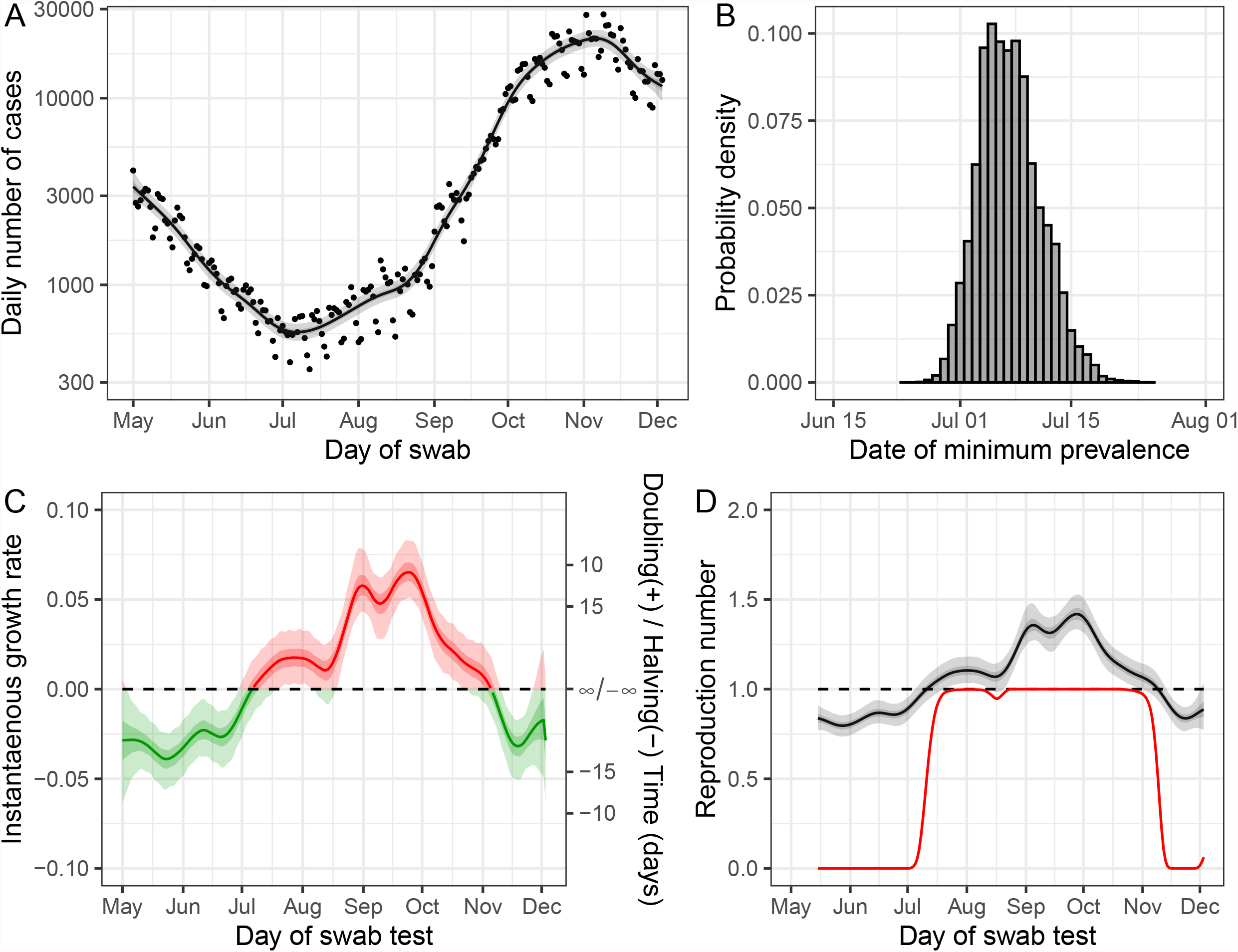
Model fit to publicly available case data. (A) Fit of the Bayesian P-spline model to publicly available case data. The central estimate (solid line) is shown with 50% (dark grey shaded region) and 95% (light grey shaded region) credible intervals and with raw data (points). (B) Posterior probability distribution for the date at which case numbers were at its lowest in England, estimated from the Bayesian P-spline model. (C) The instantaneous growth rate over the study period as inferred from the Bayesian P-spline model. The Y axis on the right shows the corresponding doubling/ halving time corresponding to the growth rate on the left Y axis. The dotted line shows where growth rate = 0 and so the point of transition between epidemic growth (red region) and decline (green region). The central estimate (solid line) is shown with 95% (light shaded region) and 50% (dark shaded region) credible intervals. (D) The rolling two week average instantaneous Reproduction number over the duration of the study calculated from the prevalence estimates of the Bayesian P-spline model. The central estimate (solid line) is shown with 95% (light grey shaded region) and 50% (light grey shaded region) credible intervals. Also shown is the probability that the reproduction number is greater than 1 over time (red line). The dashed line shows *R*_*t*_=1, the points of transition between epidemic growth and decline.

## 4 Discussion

We have developed a Bayesian P-spline model widely applicable to either prevalence or incidence epidemic time-series data and demonstrated its utility by fitting it to the first 7 rounds of the REACT-1 study. This period covered the end of the first national lockdown in England [30] and the gradual easing of restrictions. The Bayesian P-spline model allows smooth estimates of prevalence over time (even in the periods of no data between rounds) that limits over-fitting through the inclusion of a second-order random-walk prior distribution, and concurrently avoids under-fitting through the inclusion of a high enough density of basis splines. From these prevalence estimates, average growth rates over a period of time (including daily for instantaneous growth rate), and average reproduction numbers over a period of time can straightforwardly be calculated. We see that average behaviour over a single round is broadly consistent with estimates based on a simple exponential model with overlapping confidence intervals, though point estimates can differ substantially. The model has effectively included a prior distribution based on the previous and subsequent rounds’ estimated prevalence and trend. However, the Bayesian P-spline model is not limited to looking at average behaviour over the duration of a single round (a few weeks) and allows trends at finer timescales to also be quantified.

The REACT-1 testing procedure has its own set of limitations [12], for example it measures prevalence, which will be affected by individuals who remain positive for longer periods of time [31], and not incidence. However, it is relatively unbiased compared to traditional testing systems [11] and thus allows a representative picture of the true nature of the epidemic within the population of England to be obtained. From modelled prevalence estimates we calculated that the date of minimum prevalence came in mid-July to early-August 2020. This estimate was later than estimates obtained using Public Health England Pillar 1 and Pillar 2 data [10] but can be explained by increasing testing capacity obscuring underlying trends [11], especially as Pillar 2 mass testing of the community only began on July 14 2020. Prevalence was seen to increase temporarily in late-May/early-June 2020 with a high probability of the instantaneous growth rate being positive for two to three weeks. This is possibly explained by the easing of restrictions at the beginning of June 2020 [30] leading to a temporary increase in contact rates. The same temporary increase of growth rate was initially observed in the Office for National Statistics’ Covid-19 Infection Survey (ONS CIS) [32], but this increase was not present in the ONS CIS’ s subsequent modelled prevalence estimates [33].

Trends in prevalence over time were assessed by calculating the rolling 2-week average *R*_*t*_. *R*_*t*_ was found to increase temporarily above 1 in early June 2020 returning to a relative minimum at the end of June 2020. *R*_*t*_ then gradually increased over the next two months, reflecting the slow easing of restrictions including hospitality business (such as restaurants) opening from the 4 July 2020 [34], with financial incentives offered to encourage people to eat out in August 2020 [35]. We observed a plateau in our estimate of *R*_*t*_ at around 1.5 at the beginning of September 2020 that decreased to approximately 1.2 for the second half of September. This suggests that NPI’ s introduced in September including the “rule of 6” (limiting social interactions to no more than 6 people) on the 14 September 2020 [4], and 10pm closures of hospitality businesses from the 24 September 2020 [36] were only mildly effective at reducing transmission. During October 2020, *R*_*t*_ began to increase, peaking at approximately 1.4 before decreasing temporarily in late-October, perhaps due to the week-long school “half-term” holiday in late-October temporarily reducing contacts. During November 2020 and the implementation of a second national lockdown in England [37] *R*_*t*_ once more fell below 1, but this did not last for the entirety of November with *R*_*t*_ increasing in late November, in line with the timing of the introduction and subsequent growth of the Alpha variant in England [38]. It is worth noting that our estimates of *R*_*t*_ are dependent on the particular parameter estimates used for the generation time and so cannot be compared directly in magnitude with other estimates (unless they have used similar parameters). However, trends in time can be compared. Estimates published by the Scientific Advisory Group for Emergencies were not comparable because they were based on a variety of different data sources and reflected levels of transmission 2-4 weeks prior to their publication date [39].

The model applied to epidemic time-series data by the ONS CIS is comparable to ours [32]. It has the advantage over our model of including region and age as random effects, but the prior distribution it uses, penalises the magnitude of the link function of prevalence, which likely favours a constant prevalence. As in all Bayesian models it is important that the chosen prior distribution does not incorrectly influence the posterior distribution. We demonstrated how the use of a first-order random-walk prior distribution would improperly favour a constant prevalence in the absence of data. We avoided this problem by instead penalising the 1st derivative of the link function, which favours a constant growth rate. Although this is the most applicable prior distribution for the model, penalising the 1st derivative of the link function has its own limitations. Firstly, changes in growth rate are likely to be smoothed over time, whereas many policy changes are introduced overnight and so a step change in growth rate is probably more realistic. Additionally, during an epidemic with no changes in policy or behaviour, growth rate is only approximately constant over small timescales due to a decreasing proportion of susceptible individuals over longer timescales. We do not expect this to have been a problem for the current data set as a previous study estimated that only 5.56 percent of people tested positive for IgG antibodies to SARS-CoV-2 during October 27 - November 10 2020 [40], suggesting there had been a limited number of infections prior to this period.

The Bayesian P-spline model has clear advantages in estimating the underlying temporal patterns of prevalence when there are large gaps in time-series data, interpolating between periods of missing data. It can further be applied to more general epidemic time-series as we have demonstrated with the publicly available case data. It is a particularly useful tool when the growth rate can change over smaller timescales, as in the current pandemic, as the model has been shown to be sensitive to these changes whereas a more simplistic exponential model can miss these fine details. Furthermore, with the large amounts of data available on variants of infection [38, 41–43] the model could easily be applied to the relative proportion of two variants in order to test for a changing fitness advantage over time. Given that growth rate and *R*_*t*_ can be estimated from the prevalence estimates of the model it also has potential use as a forecasting tool, though more work would have to be completed looking at the sensitivity of the final growth rate estimates and optimising the window over which growth rate, and R, are estimated in order to improve predictive power.

## 5 Conclusion

In summary, we have developed a versatile model for use with epidemic time series data and applied it to the first 7 rounds of the REACT-1 study. The REACT-1 data, due to its random sampling procedure, is relatively unbiased compared to other sources of epidemic data in England over this same period. Through the application of the Bayesian P-spline model to the REACT-1 data we have been able to infer the state of the SARS-CoV-2 pandemic within England from 1 May 2020 to 3 December 2020, a period that saw numerous changes in restrictions and testing procedures. The trends we report contrast with the reported trends based on other data sets due to potential sources of bias. This study highlights the importance of not only obtaining relatively unbiased data, such as the REACT-1 data, but also in pairing it with statistically robust methods in order to effectively track the within-country dynamics of a pandemic such as the current SARS-CoV-2 pandemic.

## Data Availability

Access to individual level REACT-1 data is restricted due to ethical and security considerations. Summary statistics and data, including the daily number of positive tests and daily total number of tests, are available at https://github.com/mrc-ide/reactidd/tree/master/inst/extdata. Additional summary statistics and results from the REACT-1 programme are also available at https://www.imperial.ac.uk/medicine/research-and-impact/groups/react-study/real-time-assessment-of-community-transmission-findings/.
REACT-1 study materials are available for each round at https://www.imperial.ac.uk/medicine/research-and-impact/groups/react-study/react-1-study-materials/.

## Supplementary information

Supplementary tables are available in the supporting document ‘ SupplementaryTables.xlsx’. Supplementary figures are available in the supporting document ‘ SupplementaryFigures.docx’.

## Acknowledgments

SR, CAD acknowledge support: MRC Centre for Global Infectious Disease Analysis, National Institute for Health Research (NIHR) Health Protection Research Unit (HPRU), Wellcome Trust (200861/Z/16/Z, 200187/Z/15/Z), and Centres for Disease Control and Prevention (US, U01CK0005-01-02). GC is supported by an NIHR Professorship. PE is Director of the MRC Centre for Environment and Health (MR/L01341X/1, MR/S019669/1). PE acknowledges support from Health Data Research UK (HDR UK); the NIHR Imperial Biomedical Research Centre; NIHR HPRUs in Chemical and Radiation Threats and Hazards, and Environmental Exposures and Health; the British Heart Foundation Centre for Research Excellence at Imperial College London (RE/18/4/34215); and the UK Dementia Research Institute at Imperial (MC PC 17114). We thank The Huo Family Foundation for their support of our work on COVID-19. We thank key collaborators on this work – Ipsos MORI: Kelly Beaver, Sam Clemens, Gary Welch, Nicholas Gilby, and Kelly Ward; Institute of Global Health Innovation at Imperial College: Gianluca Fontana, Dr Hutan Ashrafian, Sutha Satkunarajah and Lenny Naar; North West London Pathology and Public Health England for help in calibration of the laboratory analyses; NHS Digital for access to the NHS register; and the Department of Health and Social Care for logistic support. SR acknowledges helpful discussion with attendees of meetings of the UK Government Office for Science (GO-Science) Scientific Pandemic Influenza – Modelling (SPI-M) committee.

## Declarations

### Data availability

Access to individual level REACT-1 data is restricted due to ethical and security considerations. Summary statistics and data, including the daily number of positive tests and daily total number of tests, are available at https://github.com/mrc-ide/reactidd/tree/master/inst/extdata. Additional summary statistics and results from the REACT-1 programme are also available at https://www.imperial.ac.uk/medicine/research-and-impact/groups/react-study/real-time-assessment-of-community-transmission-findings/. REACT-1 study materials are available for each round at https://www.imperial.ac.uk/medicine/research-and-impact/groups/react-study/react-1-study-materials/.

### Code availability

All code is available in the reactidd R package available at https://github.com/mrc-ide/reactidd/.

### Authors’ contributions

S.R. is the corresponding author. P.E. and S.R. conceived the study. O.E. developed the statistical methodology and performed the statistical analyses. O.E., K.E.C.A., C.E.W. and H.W. curated the data. C.A., D.A., C.A.D., G.C., W.B., H.Ward and A.D. provided study oversight and results interpretation. A.D. and P.E. obtained funding. O.E. and S.R. wrote the manuscript. All authors revised the manuscript for important intellectual content and approved the submission of the manuscript. P.E. and S.R. had full access to the data and take responsibility for the integrity of the data and the accuracy of the data analysis and for the decision to submit for publication.

