## Supplementary Figures for "Appropriately smoothing prevalence data to inform estimates of growth rate and reproduction number"

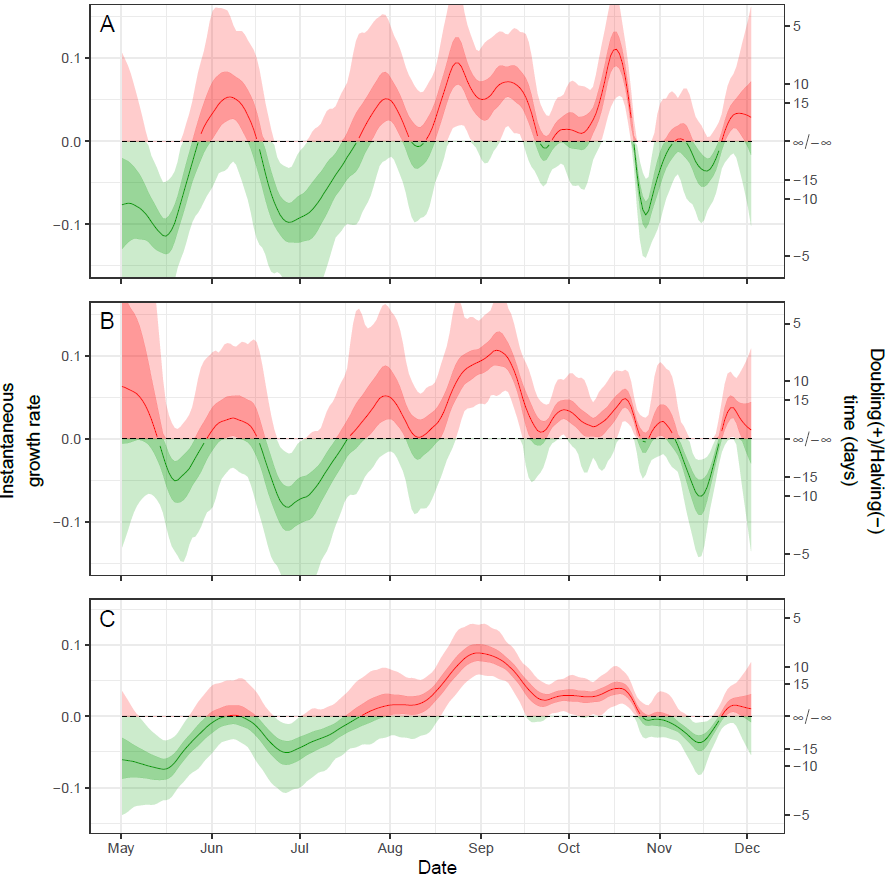

**Supplementary figure 1: Sensitivity analysis for instantaneous growth rate estimates** The instantaneous growth rate over the study period as inferred from the Bayesian P-Spline model fit to subsets of data only including positive individuals that were (A) asymptomatic, (B) had both N- and E-gene detected, and (C) were classified as positive with a lower threshold N-gene Ct-value of 35 defining positivity. The Y axis on the right shows the corresponding doubling/ halving time corresponding to the growth rate on the left Y axis. The dotted line shows where growth rate = 0 and so the point of transition between epidemic growth (red shaded region) and decline (green shaded region). The central estimates (solid line) are shown with 95% (light shaded regions) and 50% (dark shaded regions) credible intervals.

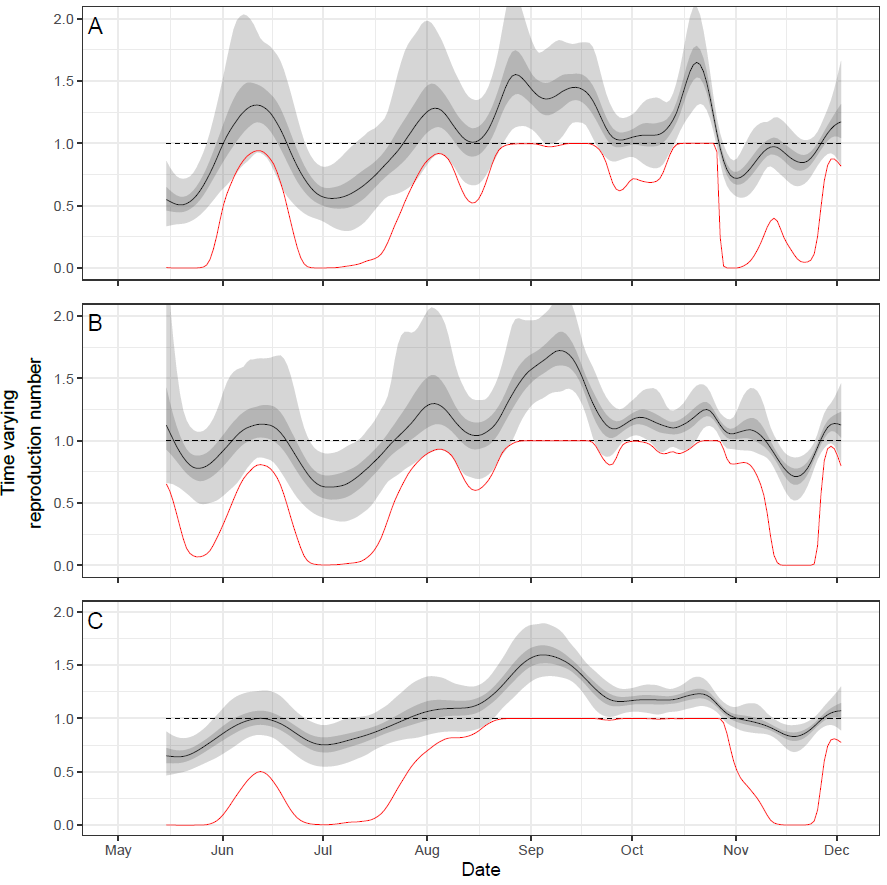

**Supplementary figure 2: Sensitivity analysis for continuous R estimates** The rolling two week average Reproduction number over the duration of the study calculated from the prevalence estimates of the Bayesian P-Spline model fit to subsets of data only including positive individuals that were (A) asymptomatic, (B) had both N- and E-gene detected, and (C) were classified as positive with a lower threshold N-gene Ct-value of 35 defining positivity. The central estimates (solid black line) are shown with 95% (light grey shaded regions) and 50% (dark grey shaded regions) credible intervals. Also shown is the probability that the reproduction number is greater than 1 over time (red line). The dashed line shows $R_{t}=1$, the points of transition between epidemic growth and decline.

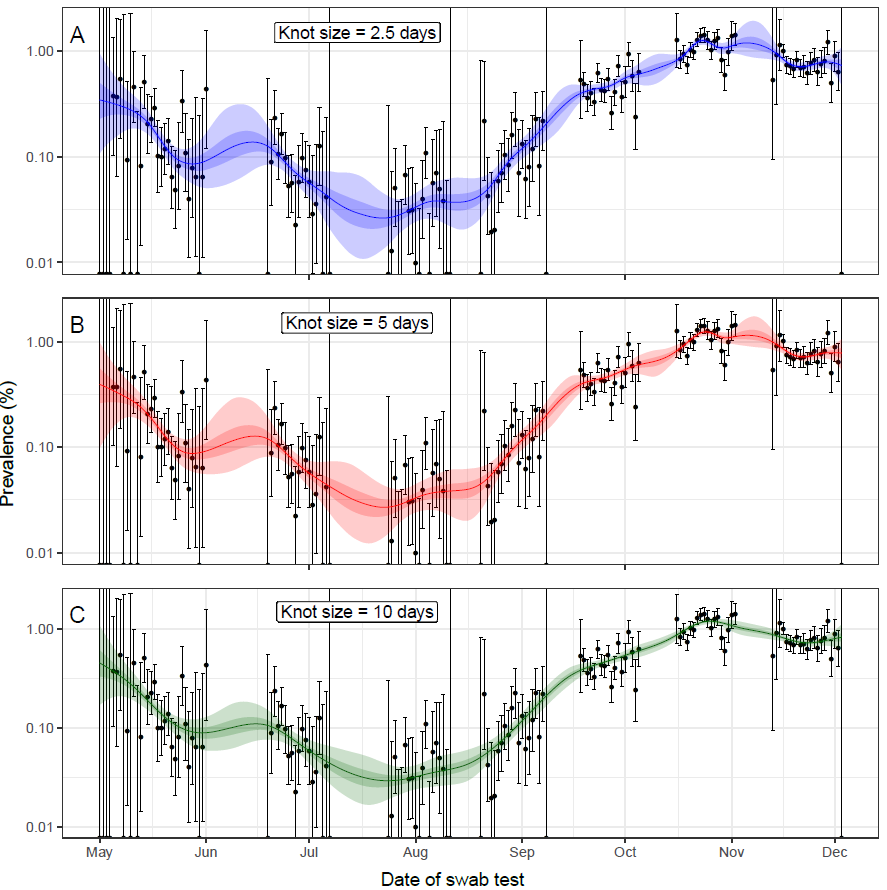

**Supplementary figure 3: Comparison of fits using different knot sizes** Bayesian P-spline model fit to the first 7 rounds of REACT-1 for different target knot sizes; target knot size = 2.5 (A, blue), target knot size = 5 (B, red), target knot size = 10 (C, green). Central estimates (solid line) are shown with 50% (dark shaded region) and 95% (light shaded regions) credible intervals. Daily prevalence estimates (points) are shown with 95% confidence intervals (error bars).

**Supplementary tables**

Both supplementary tables are available in the accompanying spreadsheet ‘SupplementaryTables.xlsx’

**Supplementary table 1: Number of tests completed** Total number of swabs, positive swabs and the number that had a date available for each round of the REACT-1 study.

**Supplementary Table 2: Estimates of average growth rates over fixed periods** Estimated growth rates and their corresponding R and doubling/halving times for fixed periods of time calculated for the exponential model and the Bayesian P-spline models
